# Microvascular Remodeling and Endothelial Dysfunction Across Post-COVID-19 and ME/CFS: Insights from the All Eyes on PCS Study

**DOI:** 10.64898/2026.01.22.26344661

**Authors:** Timon Wallraven, Roman Günthner, Isabelle Lethen, Andrea Ribeiro, Maciej Lech, Frederike Cosima Oertel, Lukas G. Reeß, Bernhard Haller, Lukas Streese, Henner Hanssen, Michael Wunderle, Christoph Schmaderer

**Author notes:** Corresponding authors: T. Wallraven; C. Schmaderer : Ismaninger Str. 22, 81675 München, Deutschland, 0049 89/4140 5231. These authors contributed equally as senior authors (Michael Wunderle, Christoph Schmaderer).

## Abstract

**Background:** Post-viral diseases, including post-COVID-19 syndrome (PCS) and myalgic encephalomyelitis/chronic fatigue syndrome (ME/CFS), cause substantial long-term morbidity. Persistent cardiovascular (CV) risk after acute infection highlights the need for accessible tools to quantify microvascular health.

**Methods:** All Eyes on PCS is a prospective, observational study investigating the retinal microcirculation using retinal vessel analysis (RVA). We compared RVA parameters in 102 PCS patients with 204 age- and sex-matched healthy controls (HC, matched from n = 303). Secondary matched analyses included never infected controls (NI, n = 96), recovered individuals (n = 102), PCS patients, and ME/CFS patients (n = 62). Laboratory variables, circulating markers of endothelial dysfunction (ED) and inflammation were compared between cohorts and their associations with RVA parameters were examined.

**Results:** Compared with HC, PCS patients showed reduced venular flicker-induced dilation (3.7 ± 2.2% vs. 4.5 ± 2.7%, p = 0.005), narrow retinal arterioles (CRAE, 178.3 ± 15.5 µm vs. 183.3 ± 15.9 µm, p = 0.009), and lower arteriolar-to-venular ratio (0.83 ± 0.06 vs. 0.86 ± 0.07, p = 0.004). Findings persisted after adjustment for CV factors and remained evident in an extended secondary matched analysis across NI, recovered, and PCS patients. ME/CFS patients showed the most pronounced alterations. PCS severity correlated with lower AVR (r = -0.21, p = 0.037) and reduced arteriolar FID (r = -0.21, p = 0.039), particularly for neurocognitive symptoms. IL-6, ICAM-1 and VCAM-1 were elevated in PCS and ME/CFS and lower AVR correlated with inflammatory and iron-related markers (all adjusted p < 0.01). A combined model discriminated ME/CFS patients with good accuracy (AUC = 0.80).

**Conclusions:** PCS is associated with persistent ED, most pronounced in ME/CFS patients and linked to symptom severity and ongoing inflammation. RVA may provide a noninvasive, readout of ED in post-viral syndromes.

**Trial Registration:** The All Eyes on PCS Study has previously been registered at ClinicalTrials.gov (NCT05635552).

**Novelty and Significance:** *What is known?:*

- PCS and ME/CFS are associated with persistent endothelial dysfunction and increased long-term cardiovascular risk.
- Neurocognitive symptoms in post-viral syndromes have been linked to impaired neurovascular coupling.
- Retinal vessel analysis provides a validated, non-invasive readout of systemic and cerebral microvascular health.

*What new information does this article contribute?:*

- PCS is characterized by persistent functional and structural retinal microvascular dysfunction
- Retinal endothelial dysfunction scales continuously with post-viral disease severity and is most pronounced in patients fulfilling ME/CFS criteria.
- Retinal microvascular alterations are linked to inflammatory–endothelial activation and iron dysregulation, identifying a biologically coherent vascular phenotype. This study provides the first comprehensive human in vivo assessment of retinal microvascular structure and function across the full post-COVID-19 spectrum, from never infected controls to recovered individuals, PCS patients, and those fulfilling ME/CFS criteria. Using retinal vessel analysis as a surrogate of neurovascular and endothelial function, we demonstrate that endothelial dysfunction persists in patients with ongoing post-viral symptomatology. Retinal venular flicker-induced dilation, arteriolar caliber, and autoregulatory capacity decline progressively with increasing clinical severity, indicating a dose-response relationship between microvascular injury and post-infectious disease burden. Importantly, these vascular alterations are linked to sustained inflammatory and endothelial activation and to disturbances in iron homeostasis, indicating an inflammatory-endothelial axis rather than isolated cardiovascular risk. By integrating microvascular phenotyping with symptom profiles and circulating biomarkers, this work identifies retinal endothelial dysfunction as a mechanistically informative and clinically accessible marker of post-viral disease severity. These findings advance understanding of post-infectious vascular pathology and provide a translational framework for biological stratification and risk assessment in PCS and ME/CFS.

## Background

Post-COVID-19 syndrome (PCS), also referred to as long COVID, remains a major public health challenge, frequently resulting in prolonged morbidity, work incapacity, and a substantial socioeconomic burden [1–4]. Although multiple terms and definitions are used by different authorities, PCS is commonly defined as the persistence or development of new symptoms three months after the initial onset of SARS-CoV-2 infection, lasting for at least two months and not attributable to alternative diagnoses. While many individuals recover fully from acute infection, a considerable subset experience persistent, multisystem symptoms, including fatigue, cognitive impairment, and exercise intolerance, that may persist for months to years [5, 6]. A subset of PCS patients fulfills diagnostic criteria for myalgic encephalomyelitis/chronic fatigue syndrome (ME/CFS), characterized by profound fatigue and post-exertional malaise (PEM) [7, 8].

Multiple, potentially interacting mechanisms, include viral persistence, immune dysregulation, autoimmunity, metabolic and mitochondrial dysfunction, muscle and neurovascular alterations, and persistent endothelial dysfunction (ED) [6, 9–13]. While these processes contribute to marked biological heterogeneity of post-viral syndromes such as PCS and ME/CFS, they may converge on shared downstream alterations at the level of endothelial function [14–16]. Subsequently, elevated blood biomarkers of ED, including intercellular adhesion molecule-1 (ICAM-1) and vascular cell adhesion molecule-1 (VCAM-1), vascular endothelial growth factor (VEGF), pro-inflammatory chemokines and cytokines including C-C motif chemokine ligand-5 (CCL5), interleukin-6 (IL-6), monocyte chemoattractant protein-1 (MCP-1), and C-X-C motif chemokine ligand-10 (CXCL10), as well as procoagulant markers such as von Willebrand factor (vWF), D-dimers, and thrombocytes, have been reported in acute COVID-19, post-COVID-19 syndrome (PCS), and myalgic encephalomyelitis/chronic fatigue syndrome (ME/CFS) [17–20].

SARS-CoV-2 infection confers a sustained increase in cardiovascular (CV) risk extending beyond the acute phase, even after mild infection, supporting the concept of persistent vascular injury and accelerated vascular ageing as a central long-term consequence of COVID-19 [21–23]. Consequently, persistent macrovascular and microvascular dysfunction assessed by flow-mediated dilation (FMD), pulse-wave velocity (PWV), post-occlusive reactive hyperemia (POH) and optical coherence tomography angiography (OCT-A) have been reported in patients with ongoing symptoms [16, 21, 24–26].

Notably, neurocognitive symptoms such as fatigue, impaired concentration, and reduced executive function have been linked to neurovascular involvement and disturbed cerebral perfusion in post-viral diseases, suggesting that ED may contribute to central nervous system manifestations [10, 27]. In this context, the retina represents an accessible surrogate of cerebral microvascular function. The retinal microvasculature shares embryological origin, anatomical features, and autoregulatory mechanisms with the cerebral microcirculation, and retinal flicker-induced vasodilation (FID) reflects neurovascular (NV) coupling mediated by NO-dependent endothelial signaling [28, 29].

Retinal vessel analysis (RVA), including static retinal vessel analysis (SVA) and dynamic retinal vessel analysis (DVA), provide a window into systemic microvascular health [30]. SVA measures retinal vessel calibers, including the central retinal arteriolar equivalent (CRAE) and central retinal venular equivalent (CRVE), which reflect microvascular structural integrity, as well as the arteriolar-to-venular ratio (AVR), calculated as CRAE divided by CRVE, which reflects retinal microvascular autoregulatory function. Changes in SVA parameters are indicative of systemic vascular health and have been validated as predictors of CV outcomes in large cohort studies [31–33].

DVA non-invasively quantifies FID of retinal arterioles (aFID) and venules (vFID) as the percentage change from baseline diameter in response to flickering light stimulation. In a cohort of patients undergoing dialysis, we previously demonstrated that reduced vFID is an independent predictor of all-cause mortality [34]. Retinal FID has been shown to correlate with dynamic cerebral hemodynamic changes assessed by functional MRI, highlighting RVA as a non-invasive approach to probe neurovascular function relevant to cognition [35].

Despite growing evidence for objective biological correlates of persistent post-viral symptoms, clinical management remains largely supportive, and accessible standardized biomarkers for diagnosis and stratification are lacking [6, 19]. Comprehensive case-control studies directly comparing endothelial health across never infected (NI) controls, SARS-CoV-2-recovered (recovered) individuals, PCS patients, and ME/CFS patients are missing. Additionally, it also remains unclear whether the degree of ED correlates with symptom severity or inflammatory status in PCS. Therefore, the primary aim of this study was to compare RVA parameters, as a non-invasive measure of microvascular endothelial health, between patients with PCS and matched healthy individuals. As secondary objectives, we compared RVA parameters across NI, recovered, PCS patients, and PCS patients fulfilling criteria ME/CFS and examined whether retinal ED was associated with patient-reported outcome measures (PROMs) and circulating markers of ED and chronic inflammation.

## Methods

### Study design and cohort

This study is based on the previously published “All Eyes on PCS” protocol, a prospective, observational, single-center study designed to examine the retinal microvasculature in PCS patients [36]. The study protocol was approved by the Ethics Committee of the Technical University of Munich, TUM School of Medicine and Health, Klinikum Rechts der Isar (Approval number: 2022-317-S-SR) and is registered at ClinicalTrials.gov (NCT05635552). The study was conducted in accordance with the Declaration of Helsinki. Sample size was defined a priori in the study protocol based on power calculations conducted with the institutional department of statistics and reflects all eligible participants recruited during the study period.

Between October 2022 and September 2023, 105 PCS patients meeting the WHO definition of post-COVID-19 condition (persistent symptoms ≥3 months after confirmed SARS-CoV-2 infection, not explained by an alternative diagnosis) were included in the study [5]. Most participants (72.4%, 76/105) were recruited through social media with the help of the patient organization “lost voices”, while the remaining (27.6%, 29/105) were referred from the PCS outpatient clinic of the LMU. Three patients were excluded from the analysis: one due to insufficient evidence of SARS-CoV-2 infection, another because of a missing temporal link between infection and PCS symptom onset, and a third because PCS-related symptoms were absent at the time of recruitment. In addition, 105 SARS-CoV-2–recovered individuals without persistent symptoms were recruited as controls and examined between October 2022 and January 2025. Three individuals were excluded from the final analysis: one due to a diagnosis of myocarditis, one due to epilepsy precluding RVA, and one due to incomplete data. NI participants were selected from a cohort of 233 individuals with no history of prior SARS-CoV-2 infection. Of these, 227 were recruited before the COVID-19 pandemic as healthy control participants as part of the Citrate-Acetate Study (recruited between 2016 and 2018; ClinicalTrials.gov: NCT02745340) and the COMPLETE Study (recruited between January 2018 and July 2019; ClinicalTrials.gov: NCT03986892) [30].Six healthy patients were recruited after the COVID-19 pandemic who reported no prior SARS-CoV-2 infection. Detailed description of patient recruitment and clinical characterization can be found in the previously published study protocol [36].

### Clinical assessment

A standardized 78-item questionnaire (Munich COVID History Questionnaire; MuCOV) was used to collect demographic data, pre-existing conditions, medication history, and PCS-related symptoms. All participants underwent a structured clinical assessment conducted by a physician. For participants recruited via social media, an initial screening survey assessed acute SARS-CoV-2 infection characteristics and persistent PCS-typical symptoms. For both PCS and recovered participants eligibility required documented proof of prior SARS-CoV-2 obtained at least three months before study inclusion. For PCS patients the presence of PCS-typical symptoms persisting for at least two months after acute infection. The temporal relationship between infection and symptom onset was reviewed in all cases, and alternative explanations for symptoms were systematically excluded. In cases of diagnostic uncertainty, eligibility was reviewed in a bi-weekly multidisciplinary meeting involving the Chief Investigator, Principal Investigator, and the study team, with final decisions made by majority vote. PROMs were assessed: PCS Severity (PCS Severity Score [37] and COVID-19 Yorkshire Rehabilitation Scale (C19-YRS) [38]), fatigue (Fatigue Severity Scale, FSS [39]), depression (Patient Health Questionnaire-9, PHQ-9 [40]), anxiety disorders (Generalized Anxiety Disorder-7, GAD-7 [41]), health-related quality of life (EuroQol 5-Dimensions 5-Levels questionnaire (EQ5DL [42]) and ME/CFS status using the Canadian Consensus Criteria [43].

### Laboratory values

Blood sampling and standard laboratory measurements were conducted as previously described [44], with IL-6, CCL-5, CXCL10, MCP-1, VCAM-1, ICAM-1 and VEGF quantified using the Cytometric Bead Array Flex system (BD Biosciences, San Diego, US). The measurements were performed according to the manufacturer’s instructions.

### Retinal vessel analysis

RVA was conducted with the Dynamic Vessel Analyzer (DVAlight; Imedos Health GmbH, Jena, Germany), while static vessel analysis (SVA) was carried out using the Static Vessel Analyzer (Imedos Health GmbH, Jena, Germany) in combination with a TRC-NW8 non-mydriatic retinal camera (Topcon, Tokyo, Japan). A detailed description of the procedure can be found in a previous publication [44].

In short: All measurements were conducted in a quiet, temperature-controlled room. Blood pressure was assessed before and after RVA to minimize potential hemodynamic confounding. Pupillary dilation was achieved using topical mydriatics. Whenever feasible, the left eye was examined to ensure inter-individual comparability. The non-examined eye was occluded during measurements. For SVA, high-resolution fundus images centered on the optic disc (field angle 50°) were acquired. At least three high-quality images were obtained per participant. Retinal arteriolar and venular calibers were semi-automatically quantified using VesselMap software (Imedos Health GmbH, Jena, Germany) and summarized as CRAE and CRVE according to the Parr–Hubbard formula; the AVR was calculated as CRAE/CRVE. One measuring unit corresponds to 1 µm in Gullstrand’s normal eye model. Images with insufficient quality were excluded (1 measurement in PCS 1/103; 0.9%). For DVA, participants were instructed to fixate on the internal target and vessel segments (approximately 0.5-1 mm in length) were selected roughly two optic-disc diameters from the optic disc margin, preferentially in the superior-temporal quadrant (alternatively inferior-temporal if required). Diameters of the selected retinal arteriole and venule were continuously recorded for a total of 350 s. Following a baseline period, three flicker-light cycles (20 s each) were applied, each followed by a recovery period. FID was calculated as percent change relative to baseline (aFID/vFID). RVA measurements were performed by trained personnel. To ensure high measurement quality, vessel response curves were rated using a cumulative scoring approach (scale 0-5). Recordings with a total score <2.5 were re-evaluated by a second experienced observer and excluded if consensus could not be reached. For vFID and aFID, 3 measurements in PCS (3/103; 2.9%) had to be excluded.

### Data analysis

All statistical analyses were performed using R (version 4.2.1; R Foundation for Statistical Computing, Vienna, Austria) in RStudio and followed the “Strengthening the Reporting of Observational studies in Epidemiology” (STROBE) guidelines. Key packages included Matching, MatchIt, WeightIt, survey, gtsummary, ggplot2, pROC, and corrplot. Analyses were conducted using complete-case data; no imputation was performed, as missingness was low and did not show systematic patterns. Normality was assessed by visual inspection of histograms and Q-Q plots and formally tested using the Shapiro-Wilk test. Normally distributed variables are presented as mean ± standard deviation (SD), and non-normally distributed variables as median with interquartile range (Q1, Q3).

For subgroup secondary matched analyses, participants were categorized into PCS, recovered, and NI groups. As recovered participants were substantially younger and showed limited age overlap with PCS, age matching was applied only for the NI reference group, whereas comparisons involving recovered were addressed using age-adjusted and overlap-weighted sensitivity analyses (ATO). PCS patients were age- and sex-matched to NI controls using nearest-neighbor matching, with balance assessed using the MatchBalance function. Due to substantial age differences between PCS and recovered cohorts, these groups were matched on sex only. Primary analyses adjusted for age and other relevant confounders using multivariable regression models. In addition, to address residual age imbalance and limited covariate overlap, we performed overlap weighting targeting the average treatment effect in the overlap population (ATO estimand) using the WeightIt package. Weighted group comparisons for RVA parameters were performed using survey-weighted t-tests via the survey package. One eye per participant was included in the analysis, such that each observation represents a single eye from a unique individual. Unweighted group comparisons were conducted using Welch’s two-sample t-test or the Mann-Whitney U test for two-group comparisons and the Chi-square (χ²) test for categorical variables. For comparisons involving more than two groups, we used one-way ANOVA when model assumptions were met; otherwise, the Kruskal-Wallis test was applied. For ANOVA, post-hoc pairwise comparisons were conducted using Tukey’s honestly significant difference (HSD) test. For Kruskal-Wallis, post-hoc pairwise comparisons were performed using Dunn’s test with multiplicity correction. Multivariable linear regression models were fitted to adjust for established confounders of RVA parameters [45–48]. Continuous biomarkers with skewed distributions or outliers were analyzed using robust linear regression based on M-estimation. Correlation analyses were performed using Pearson’s correlation for normally distributed data and Spearman’s rank correlation for non-normally distributed data. For correlation analyses involving multiple biomarkers, p-values were adjusted using the Benjamini-Hochberg procedure. For primary, pre-specified group comparisons, no multiplicity correction was applied. Receiver operating characteristic (ROC) analyses were used to assess the discriminative ability of individual variables, with confidence intervals calculated using DeLong’s method. To evaluate whether a combination of inflammatory, metabolic, and microvascular markers improves discrimination of the ME/CFS phenotype among SARS-CoV-2-infected individuals, a multivariable logistic regression model including transferrin, ICAM-1, neutrophils, creatine kinase, IL-6 and AVR was fitted (**Supplementary Table 1**). ME/CFS status served as the dependent variable. Predicted probabilities were used to construct ROC curves and estimate the area under the curve (AUC). Please see the Major Resources Table in the Supplemental Materials for detailed information on study resources (**Supplementary Table 2**).

## Results

### Cohort characteristics

**Table 1** presents the baseline demographic and clinical characteristics of the study population, showing cohort comparison between PCS patients (n = 102, age: 41.9 ± 11.6 years, 75% female) and age- and sex-matched healthy controls (HC; matched out of 303 participants, n = 204, age: 42.9 ± 15.4 years, 75% female), which includes both NI individuals (n=117) and recovered individuals (n=87). No differences were observed in BMI or systolic blood pressure between groups. CV risk factors such as arterial hypertension (16% vs. 2.0%, p < 0.001) and hypercholesterolemia (22% vs. 5.4 %, p =0.002) were more prevalent in PCS patients and were therefore used for controlling for confounders when comparing RVA parameters. Further cohort characteristics with a detailed three-group comparison are presented in later sections.

**Table 1.** Baseline demographic and clinical characteristics of the healthy control (HC) cohort and the post-COVID-19 syndrome (PCS) cohort. Continuous variables are reported as mean ± standard deviation (SD) and categorical variables as n (%). Group comparisons were performed using Welch’s two-sample t-test for normally distributed continuous variables, the Mann–Whitney U test for non-normally distributed continuous variables, and Pearson’s χ² test for categorical variables, as appropriate. Missing values were as follows: systolic blood pressure before retinal vessel analysis (RRsyst; HC n = 12, PCS n = 7) and hypercholesterolemia (HC n = 112, PCS n = 0). Abbreviations: BMI, body mass index; RRsyst, systolic blood pressure before retinal vessel analysis (RVA). Statistical significance is indicated as *p < 0.05; **p < 0.01; **p < 0.001.

| Baseline characteristics | Healthy cohort<br>N = 204 | Post-COVID-19<br>Syndrome<br>N = 102 | p-value |
| --- | --- | --- | --- |
| Age (years), mean (±SD) | 42.9<br>($\pm 15.4$) | 41.9<br>($\pm 11.6$) | 0.50 |
| Gender (female), n(%) | 154 (75%) | 77 (75%) | >0.9 |
| BMI (kg/m<sup>2</sup>), mean (±SD) | 23.7 ($\pm 3.9$) | 24.5 ($\pm 4.8$) | 0.12 |
| RR<sub>syst</sub><sup>1</sup> (mmHg), mean (±SD) | 122.2<br>($\pm 17.1$) | 121.8 ($\pm 16.9$) | 0.80 |
| Cardiovascular Risk Factors |  |  |  |
| Arterial Hypertension, n(%) | 4 (2.0%) | 16 (16%) | <0.001*** |
| Hypercholesterolemia, n(%) | 5 (5.4%) | 22 (22%) | **0.002\*\*** |
| Smoking history, n(%) | 29 (14%) | 10 (9.8%) | 0.40 |
| Diabetes mellitus (I and II), n(%) | 0 (0%) | 1 (1.0%) | 0.70 |

### Functional and Structural Retinal Microvascular Alterations in PCS vs. Healthy Cohort

RVA parameters revealed persistent retinal ED in PCS patients compared to HC. Venular flicker-induced dilation (vFID) was reduced in PCS patients (3.7 ± 2.2% vs. 4.5 ± 2.7%, p = 0.005), indicating ongoing endothelial dysfunction. Arteriolar flicker-induced dilation (aFID) was numerically lower in PCS patients (2.9 ± 2.3% vs. 3.2 ± 2.0%), though this difference was not statistically significant (p = 0.261) (**Fig. 1a and b**). PCS patients showed narrower retinal arterioles (CRAE, 178.3 ± 15.5 µm vs. 183.3 ± 15.9 µm, p = 0.009), while retinal venular diameters (CRVE) did not differ between groups (213.85 ± 16.13 µm vs. 214.02 ± 18.36 µm, p = 0.937). Consequently, PCS patients exhibited a lower AVR (0.83 ± 0.06 vs. 0.86 ± 0.07, p = 0.004) (**Fig. 1c–e**). To assess potential confounding, we adjusted for arterial hypertension and hypercholesterolemia using forest plots. After adjustment, lower vFID (Standardized [Std.] β = −0.48, 95% CI −0.77 to −0.18; p = 0.002) and CRAE (Std. β = −0.39, 95% CI −0.67 to −0.12; p = 0.006) remained associated with PCS status, whereas the association with AVR was weaker (Std. β = −0.26, 95% CI −0.55 to 0.02; p = 0.073) (**Fig. 1f**). To assess whether the association between PCS status and microvascular changes differed by sex, we included a cohort-by-sex interaction term in the linear model. No significant sex-by-cohort interactions were observed for AVR, CRAE, or vFID, suggesting that retinal ED in PCS is independent of sex (**Supplementary Table 3**).

**Figure 1.**
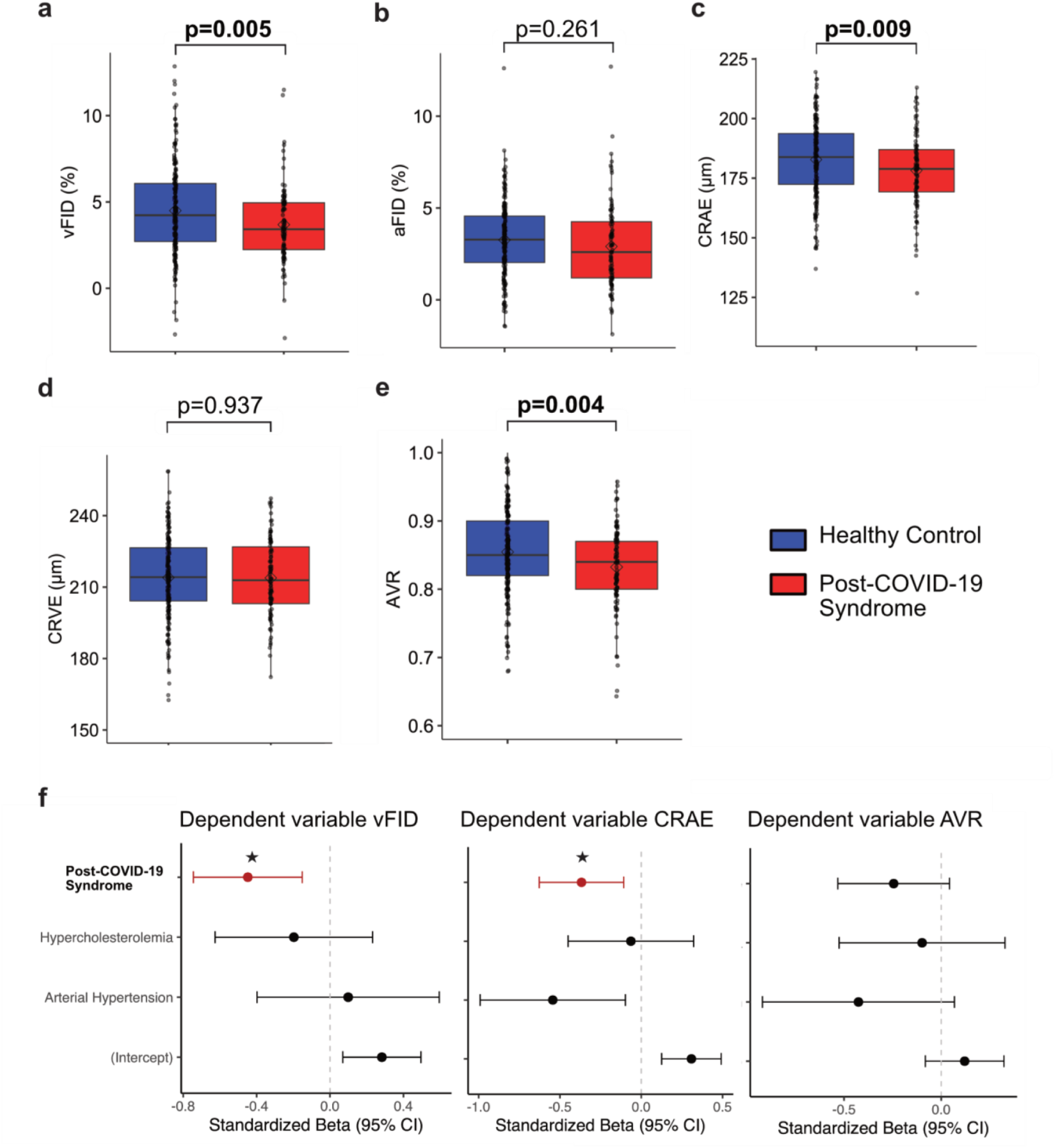
Retinal microvascular parameters in PCS patients and HC. Box plots show dynamic retinal vessel analysis (DVA) parameters: venular flicker-induced dilation (vFID; PCS n = 100, HC n = 195; a) and arteriolar flicker-induced dilation (aFID; PCS n = 100, HC n = 195; b), and static retinal vessel analysis (SVA) parameters: central retinal venular equivalent (CRVE; c), central retinal arteriolar equivalent (CRAE; d), and arteriolar-to-venular ratio (AVR; PCS n = 102, HC n = 199; e) in age- and sex-matched PCS patients (red) and healthy controls (blue). Box plots indicate the median (horizontal line) and mean (black dot). Each data point represents one eye from one individual (one eye analyzed per participant). Group comparisons were performed using the Mann-Whitney U test for non-normally distributed variables and Welch’s two-sample t-test for normally distributed variables. Forest plots (f) display standardized β coefficients with 95% confidence intervals from multivariable linear regression models assessing the independent association of PCS status with vFID, CRAE, and AVR, adjusted for arterial hypertension and hypercholesterolemia. Statistical significance is indicated as *p < 0.05.

### Cohort Characteristics for PCS patients, Recovered, and Never Infected individuals

An extended three-group comparison across age- and sex-matched NI (n = 96), sex-matched recovered (n = 102), and PCS patients (n = 102) is presented in **Supplementary Table 4**. Age differed between groups (p < 0.001), with recovered individuals being younger (33.3 ± 11.4 years, 72% female) than both NI (43.0 ± 14.5 years, 76% female) and PCS patients (41.9 ± 11.6 years, 75% female). Systolic blood pressure before RVA varied between cohorts (p < 0.05), with recovered participants showing the lowest values. CV risk such as arterial hypertension was present in 16% of PCS patients compared with 4.9% of recovered and 0% of NI individuals (p < 0.001), and hypercholesterolemia was more common in PCS than in recovered (22% vs. 5.9%).

Acute SARS-CoV-2 infection characteristics showed differences: PCS patients experienced more severe acute disease (WHO progression scale; p < 0.001) and were more frequently infected with alpha or delta variants (p = 0.004). Most recovered and PCS individuals reported a single infection (61% and 70%, respectively). PCS burden remained high, with a median symptom duration of 404 days (IQR: 291 to 575), median cumulative sick leave of 199.5 days (44.0 to 377.0), and 16% reporting occupational loss (all p < 0.001 compared with recovered). PCS symptom severity was markedly elevated (36.9 ± 10.6 vs. 1.8 ± 5.9, p < 0.001), and 61% of PCS patients fulfilled ME/CFS diagnostic criteria.

Hematologic and biochemical parameters differed between groups: PCS patients showed higher neutrophil percentages (p = 0.001), lower lymphocyte percentages (p = 0.041), and higher platelet counts (p = 0.040). Iron metabolism markers indicated elevated ferritin (p = 0.008) and reduced transferrin (p < 0.001) in PCS compared with recovered individuals, while hemoglobin was higher in NI than in both post-infection groups (p < 0.001). Lipid profiles differed across cohorts (total cholesterol: p = 0.002; LDL cholesterol: p = 0.001). Total cholesterol was lower in recovered individuals, whereas LDL cholesterol was highest in PCS patients. Creatine kinase (CK) was significantly lower in PCS (p<0.001). Immunoglobulin profiles were broadly similar, except for lower IgG3 levels in PCS patients (p = 0.021). Given the younger age of recovered participants, laboratory differences were examined in age-adjusted sensitivity analyses. In unweighted linear models including all three cohorts, hemoglobin, CRP, and leukocyte counts differed significantly between PCS patients and NI individuals (hemoglobin: standardized β = 0.20, p = 0.004; CRP: standardized β = −0.41, p < 0.001; leukocytes: standardized β = −0.14, p = 0.028), with no difference between recovered and PCS. In complementary age-balanced sensitivity analyses using overlap weighting (ATO), differences in neutrophil counts (mean difference +3.97%, CI 1.05 to 6.90; p = 0.008), transferrin (mean difference −16.57 mg/dL, CI −27.90 to −5.24; p = 0.004), and CK between PCS and recovered remained significant, while differences in lymphocytes, platelets, ferritin, lipid parameters, IgG3, and hemoglobin were attenuated (**Supplementary Table 5 and 6**).

### Functional and Structural Retinal Microvascular Alterations Across PCS, Recovered, and Never Infected

Mean venular FID (vFID) differed between the three groups (p = 0.007). In post-hoc comparisons, PCS patients showed lower vFID than recovered individuals (3.7% ± 2.2 vs. 4.8% ± 3.0, p = 0.005). No differences were observed between PCS and NI (4.1% ± 2.5, p = 0.541) or between recovered and NI participants (p = 0.101). Arteriolar FID showed a trend toward lower values in PCS compared with recovered individuals (2.9% ± 2.3 vs. 3.4% ± 2.1), but this did not reach significance (**Fig. 2a and b**).

**Figure 2.**
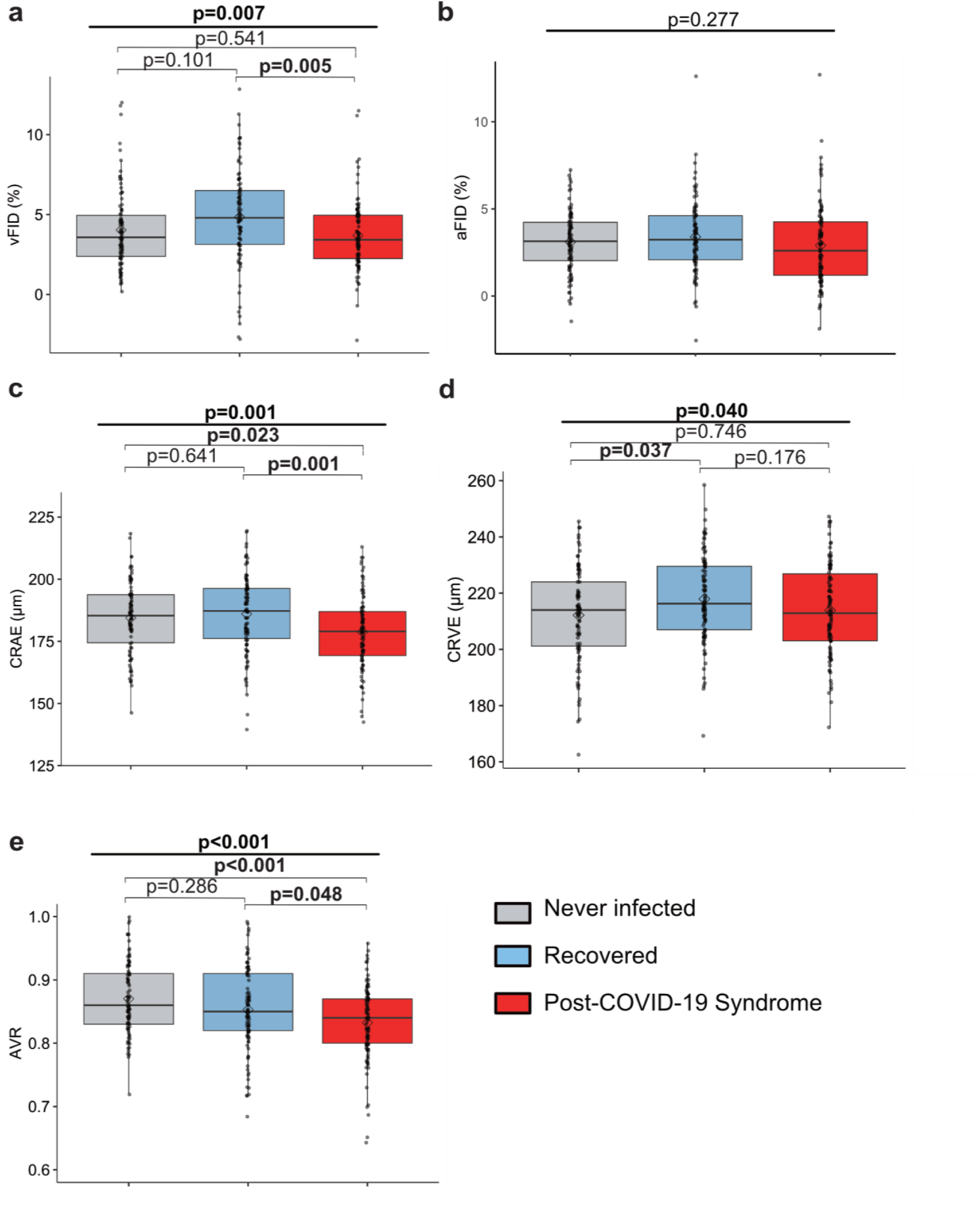
Retinal microvascular parameters across never infected, recovered, and PCS cohorts. Box plots show DVA parameters vFID and aFID (never infected n = 95; recovered n = 92; PCS n = 100; a, b), and SVA parameters CRAE (c), CRVE (d), and AVR (never infected n = 94; recovered n = 99; PCS n = 102; e). Groups are visualized as never infected (grey), recovered (light blue), and PCS (red). Box plots indicate the median (horizontal line) and mean (black dot). Each data point represents one eye from one individual (one eye analyzed per participant). Group comparisons were performed using one-way ANOVA for normally distributed variables or the Kruskal-Wallis test for non-normally distributed variables. Post-hoc pairwise comparisons were conducted using Tukey’s honestly significant difference (HSD) test for ANOVA models and Dunn’s test with multiplicity correction for Kruskal-Wallis models.

Retinal arteriolar diameters differed across groups (p = 0.001). PCS patients had narrower arterioles compared to both recovered (178.3 ± 15.5 µm vs. 186.1 ± 15.7 µm, p = 0.001) and NI (184.1 ± 15.1 µm, p = 0.023), whereas recovered and NI did not differ (p = 0.641). Retinal venular diameters also differed between groups (p = 0.040). Recovered had slightly larger venular calibers than NI (217.9 ± 15.5 µm vs. 212.2 ± 16.9 µm, p = 0.037), while differences between recovered and PCS were not significant (p = 0.176) (**Fig. 2c and d**). AVR differed between groups (p < 0.001). PCS patients exhibited lower AVR compared with both NI (0.83 ± 0.06 vs. 0.87 ± 0.06, p < 0.001) and recovered (0.86 ± 0.07, p = 0.037). (**Fig. 2 e**). Age, arterial hypertension, BMI, and systolic blood pressure before RVA measurement were included as confounders, with hypercholesterolemia added in a sensitivity model for PCS vs. recovered. After adjustment, higher vFID remained significantly associated with recovered (Std. β = 0.25, p < 0.001). Higher CRAE remained significantly associated with NI (Std. β = 0.15, p = 0.021), whereas the association with recovered (Std. β = 0.11, p = 0.105) was attenuated. For CRVE associations were no longer significant. For AVR, higher values remained significantly associated with NI (Std. β= 0.21, p = 0.002), whereas the association with recovered was weaker (Std. β= 0.10, p = 0.167) (**Supplementary Table 7**).

In ATO analysis, vFID remained significantly reduced in PCS patients compared with recovered individuals (mean difference -1.3%, CI: -2.0 to -0.5%, p = 0.001). Retinal arteriolar narrowing also persisted, with lower CRAE in PCS (mean difference -5.6 µm, CI -10.3 to -0.8 µm, p = 0.022). In contrast, the difference in AVR was attenuated and narrowly missed statistical significance (mean difference -0.02, CI -0.04 to - 0.0003, p = 0.053), indicating a small residual effect after age balancing. CRVE and aFID did not differ between groups following age adjustment. (**Supplementary Table 8**). Sensitivity analyses including hypercholesterolemia did not alter the results (not shown).

### Association of Patient-Reported Outcome Measurements with Persistent Changes of Retinal Microcirculation

PROMs, including the PCS Severity Score and the COVID-19 Yorkshire Rehabilitation Scale (C19-YRS), were associated with persistent alterations in retinal microcirculation. AVR showed a significant negative correlation with both PCS severity score (r = -0.21, p = 0.037) and C19-YRS scores (r = -0.20, p = 0.043) (**Fig 3 a and b**). Likewise, aFID was inversely correlated with the PCS severity score (r = -0.21, p = 0.039; **Fig. 3 d**), however not with C19-YRS score (**Fig. 3 e**). In regression analyses, models were adjusted for sex and BMI, as age and CV risk factors were not associated with PCS severity within this subgroup. In these models, lower aFID remained independently associated with PCS severity score, whereas the association between lower AVR and symptom severity was attenuated (**Fig. 3 c and f**). vFID, CRAE, and CRVE showed no significant association with PCS severity within the PCS cohort.

**Figure 3.**
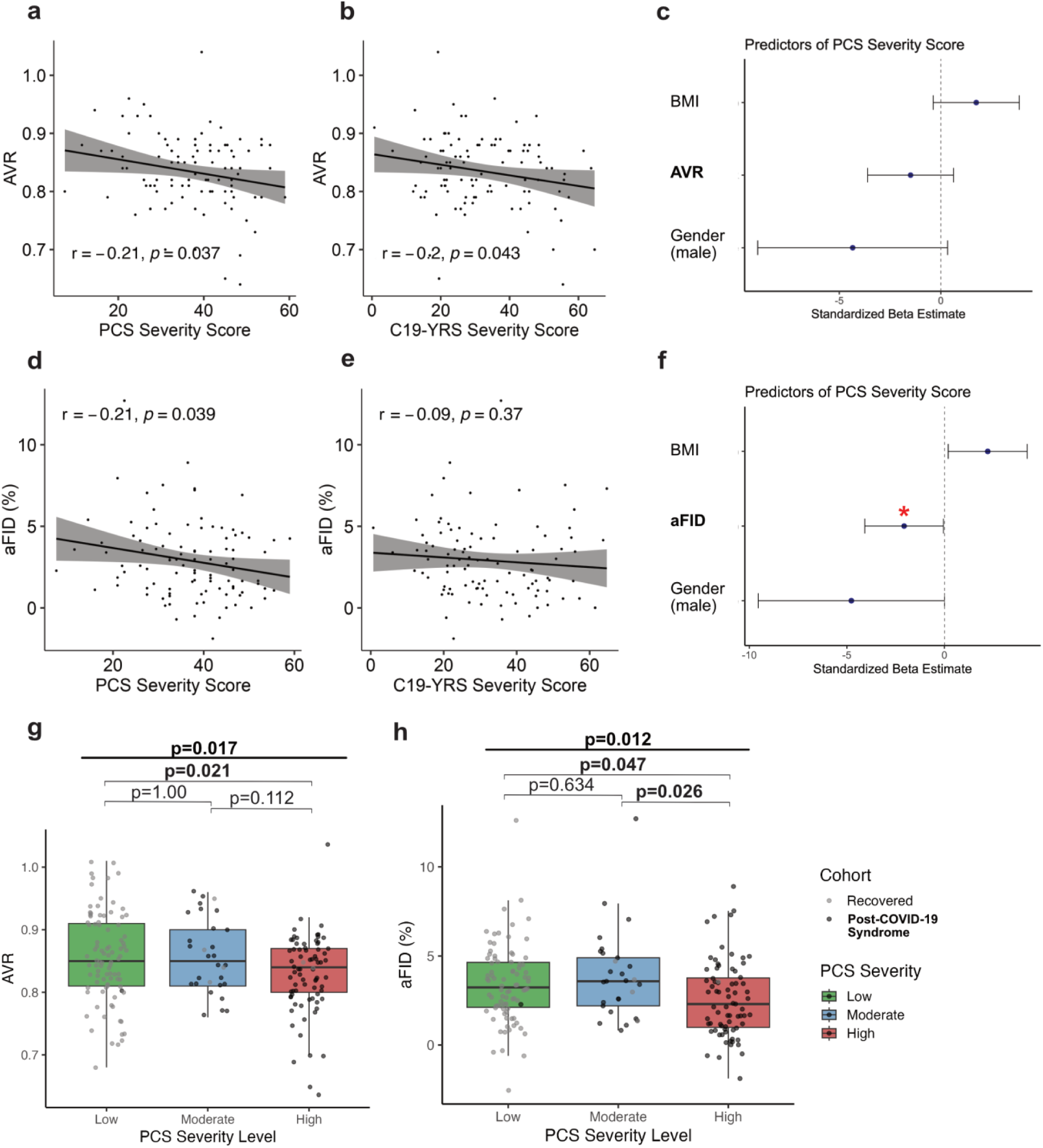
Associations between RVA parameters and PCS symptom burden. **(a–b, d–e)** Pearson correlations between RVA parameters and symptom scores in PCS patients. Panels **a–b** show correlations between AVR and the PCS Severity Score (**a**) and C19-YRS (**b**). Panels **d–e** show corresponding correlations for aFID with the PCS Severity Score (**d**) and C19-YRS (**e**). Points represent individual patients; the solid line indicates the linear regression fit and the shaded band the 95% confidence interval. Pearson correlation coefficients (r) and p-values are shown in each panel. **(c, f)** Forest plots display standardized β-coefficients with 95% confidence intervals from multivariable regression models assessing the association of AVR (**c**) or aFID (**f**) with the PCS Severity Score after adjustment for BMI and gender. Statistical significance is indicated as *p < 0.05. **(g–h)** Box plots show the distribution of AVR (**g**) and aFID (**h**) across three symptom severity groups in PCS patients (black) and recovered individuals (grey). The PCS Severity Score was categorized into low (green), moderate (blue), and high (red) severity using fixed cutoffs (<10, 10–30, >30; colors as indicated). Box plots indicate the median (horizontal line) and individual observations. Group comparisons were performed using one-way ANOVA for normally distributed variables. Post-hoc comparisons were conducted using Tukey’s HSD for ANOVA models. Each data point represents one eye from one individual (one eye analyzed per participant).

To further investigate the relationship between symptom burden and RVA parameters, all previously infected individuals (recovered and PCS) were stratified into tertiles based on PCS severity scores. The low symptom severity group predominantly comprised recovered (light grey), while the moderate and high severity groups were largely composed of PCS patients (dark grey). For AVR, a significant overall difference across severity levels was observed (p = 0.017), with post-hoc comparisons revealing significantly lower AVR values in the high vs. low severity group (p = 0.021). For aFID, the overall group difference was significant (p = 0.012), with reduced aFID observed in both the moderate vs. high (p = 0.026) and low vs. high (p = 0.047) severity comparisons (**Fig. 3 g and h**).

To further stratify which individual symptoms are most closely linked to persistent retinal ED, we compared AVR values between previously infected individuals reporting versus not reporting each of the 16 key symptoms included in the PCS severity score. AVR was significantly lower in participants reporting symptoms predominantly within the neurocognitive cluster (e.g., fatigue, concentration difficulties, headache, balance/fine motor dysfunction) and systemic/malaise-related symptoms (e.g., general malaise, hair/skin changes, gastrointestinal symptoms). In contrast, AVR did not differ significantly for symptoms primarily related to respiratory or sensory domains (e.g., chest pain, shortness of breath, cough, disturbed smell/taste) (**Table 2**).

**Table 2.**
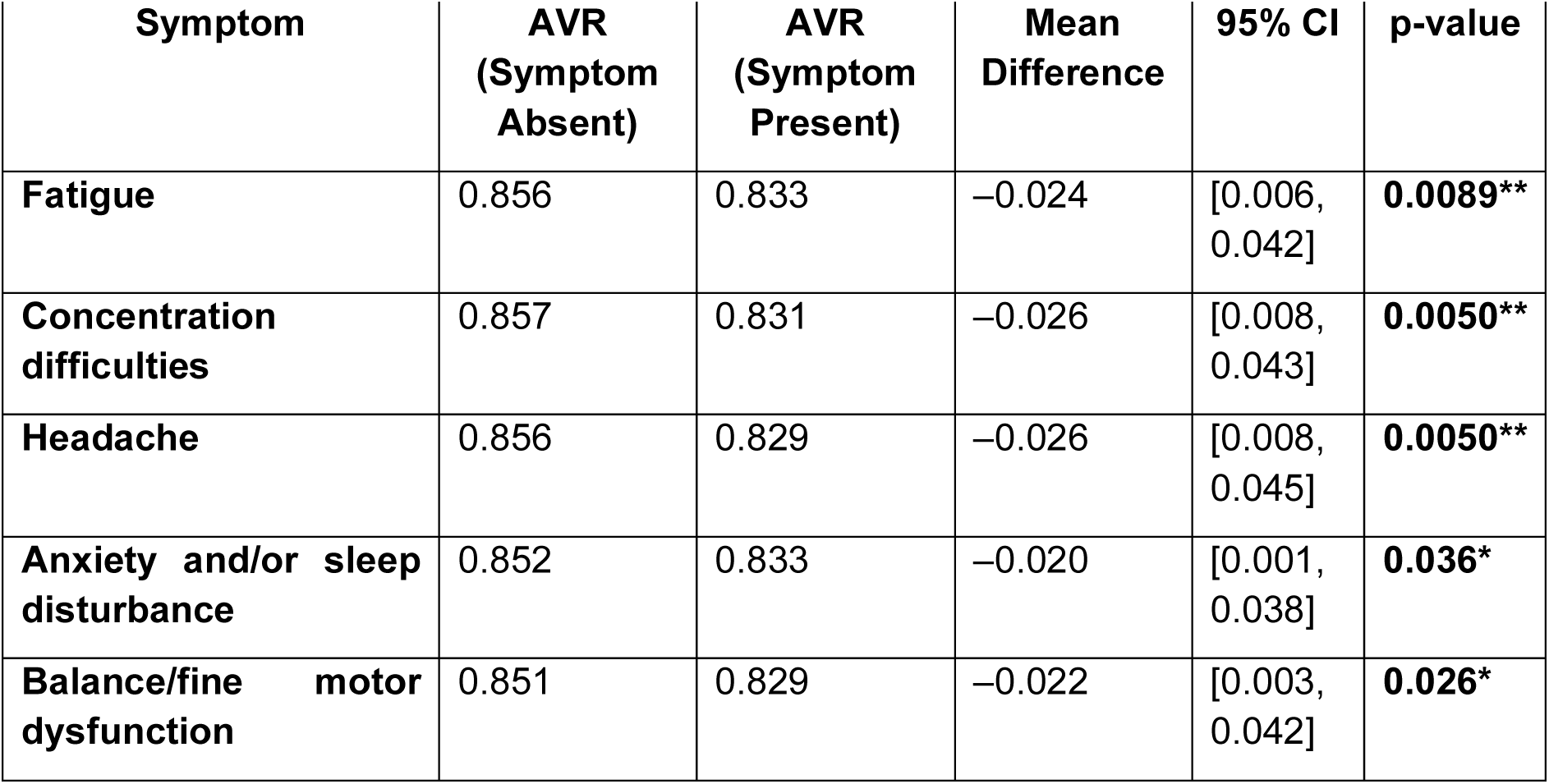
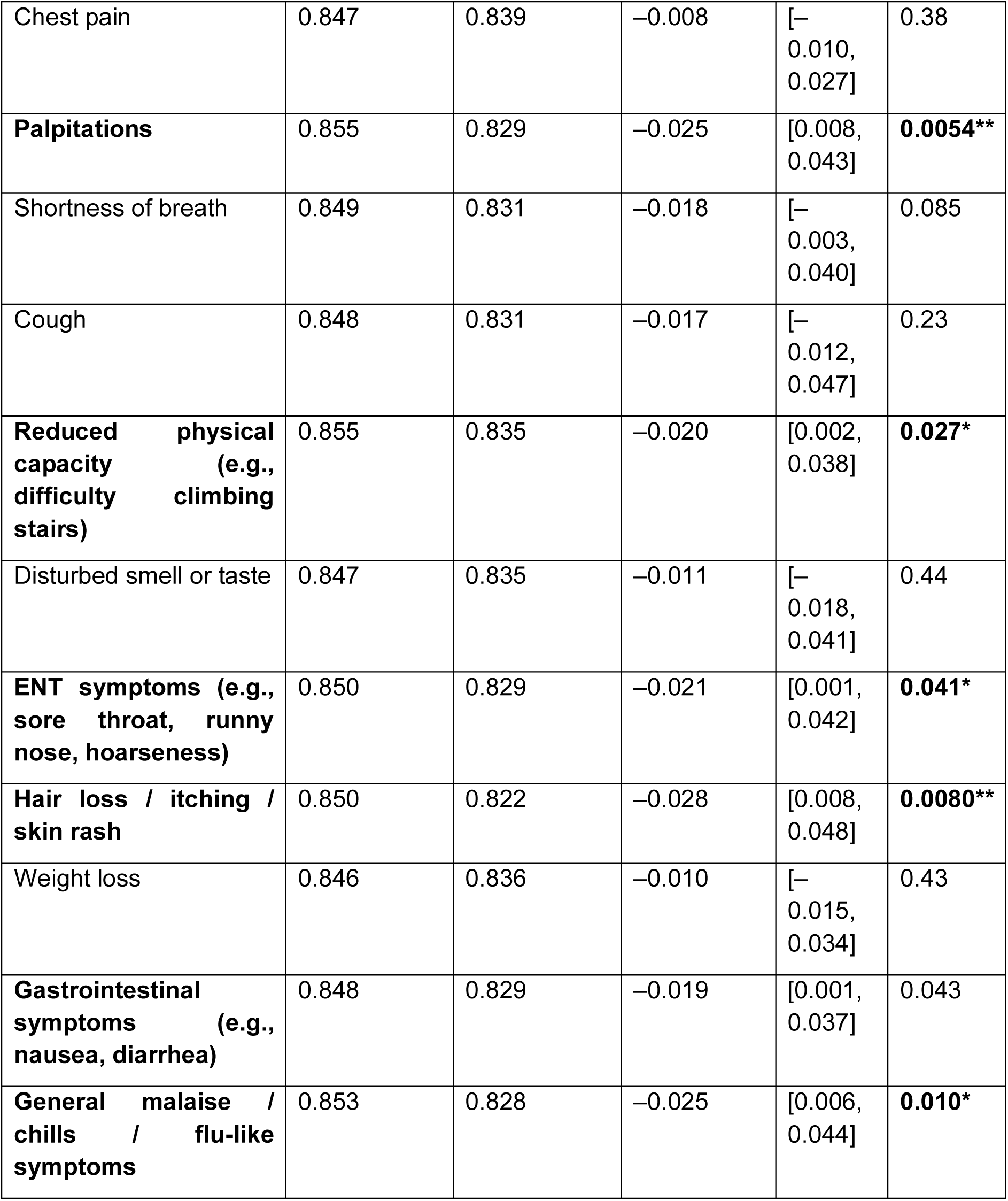
Mean AVR in participants who did not report (“Symptom absent”) versus did report (“Symptom present”) each of the 16 symptoms included in the PCS Symptom Severity Score. For each symptom, mean AVR values are shown for both groups, along with the mean difference calculated as Present minus Absent, corresponding 95% confidence intervals, and p-values from group comparisons. Symptoms are listed in the order included in the PCS Symptom Severity Score and span neurological, cardiopulmonary, sensory, and systemic domains. Group comparisons were performed using Welch’s two-sample t-test or the Mann-Whitney U test, as appropriate. Statistical significance is indicated as *p < 0.05; **p < 0.01; **p < 0.001.

### Retinal Microvascular Alterations in ME/CFS

Persistent ED has previously been demonstrated in post-viral syndromes and in patients with ME/CFS [16]. In our cohort, PCS patients fulfilling ME/CFS criteria were slightly younger (p = 0.041) and exhibited a higher overall symptom burden, as reflected by elevated C19-YRS scores (p = 0.023), PCS Severity Scores (p < 0.001), PHQ-9 scores (p = 0.008), Fatigue Severity Scale scores (p < 0.001), and reduced quality of life as measured by the EQ-5D-5L index (p = 0.010). Consistent with this, patients with ME/CFS reported more cumulative sick days (p < 0.001). CV risk factors, including hypercholesterolemia, hypertension, and diabetes, did not significantly differ between groups, nor did characteristics of the acute SARS-CoV-2 infection. Laboratory parameters were largely comparable, although CK levels were significantly lower in the ME/CFS subgroup (p = 0.009) (**Table 3**).

**Table 3.** Baseline characteristics of PCS patients stratified by ME/CFS status according to the Canadian Consensus Criteria (CCC). One patient did not complete the CCC questionnaire and was therefore excluded from the analysis. Continuous variables are reported as mean ± SD or median (IQR), depending on distribution, and categorical variables as n (%). Group comparisons were performed using Welch’s two-sample t-test for normally distributed continuous variables, the Mann-Whitney U test for non-normally distributed continuous variables, and Pearson’s χ² test for categorical variables, as appropriate. Abbreviations: BMI, body mass index; RRsyst, systolic blood pressure before retinal vessel analysis; C19-YRS, COVID-19 Yorkshire Rehabilitation Scale; PHQ-9, Patient Health Questionnaire-9; GAD-7, Generalized Anxiety Disorder-7; FSS, Fatigue Severity Scale; EQ-5D-5L, EuroQol 5-Dimension 5-Level index; ME/CFS, myalgic encephalomyelitis/chronic fatigue syndrome. Statistical significance is indicated as *p < 0.05; **p < 0.01; **p < 0.001.

| Variable | PCS without ME/CFS<br>N = 39 | PCS with ME/CFS<br>N = 62 | p-value |
| --- | --- | --- | --- |
| Age (years), mean (±SD) | 44.8 (±11.6) | 39.9 (±11.2) | **0.041\*** |
| Gender (female), n(%) | 28 (72%) | 49 (79%) | 0.6 |
| BMI ($\text{kg}/\text{m}^2$), mean (±SD) | 24.4 (±4.6) | 24.6 (±4.9) | 0.8 |
| RR<sub>syst</sub><sup>1</sup>(mmHg), mean (±SD) | 121.9 (±18.8) | 121.7 (±15.6) | >0.9 |
| *Cardiovascular Risk Factors* |  |  |  |
| Diabetes mellitus (I and II), n(%) | 1 (2.6%) | 0 (0%) | >0.9 |
| Arterial Hypertension, n(%) | 5 (13%) | 11 (18%) | 0.7 |
| Hypercholesterolemia, n(%) | 7 (18%) | 15 (24%) | 0.6 |
| Smoking history, n(%) | 4 (10%) | 6 (9.7%) | >0.9 |
| *PCS-related characteristics* |  |  |  |
| C19-YRS, mean (±SD) | 28.3 (15.1) | 35.0 (12.7) | **0.023\*** |
| PCS Severity Score, mean (±SD) | 32.0 (11.7) | 40.0 (8.6) | **<0.001\*\*\*** |
| PHQ9, mean (±SD) | 8.9 (4.7) | 11.4 (3.9) | **0.008\*\*** |
| GAD7, median (IQR) | 4.0 (2.0, 7.0) | 6.0 (3.0, 9.0) | 0.2 |
| FSS, median (IQR) | 5.4 (3.7, 6.4) | 6.3 (5.8, 6.7) | **<0.001\*\*\*** |
| EQ5DL-Index, median (IQR) | 0.8 (0.6, 0.8) | 0.5 (0.3, 0.7) | **0.010\*** |
| Duration of PCS, days, median (IQR) | 345.0 (218.0, 537.0) | 409.5 (316.5, 619.0) | 0.15 |
| Cumulative sick days, median (IQR) | 129.0 (0.0, 291.0) | 285.5 (106.0, 445.0) | **<0.001\*\*\*** |
| *Acute SARS-CoV-2 infection* |  |  |  |
| Work loss during PCS, n(%) | 4 (10%) | 12 (20%) | 0.3 |
| Number of infections, n(%) |  |  | 0.2 |
| 1 | 30 (77%) | 40 (65%) |  |
| 2 | 8 (21%) | 21 (34%) |  |
| 3 | 1 (2.6%) | 0 (0%) |  |
| 4 | 0 (0%) | 1 (1.6%) |  |
| Severity of acute infection, n(%)<br>(WHO progression scale) |  |  | 0.6 |
| 1 | 1 (2.6%) | 0 (0%) |  |
| 2 | 25 (64%) | 35 (56%) |  |
| 3 | 12 (31%) | 22 (35%) |  |
| 4 | 1 (2.6%) | 3 (4.8%) |  |
| 5 | 0 (0%) | 1 (1.6%) |  |
| 6 | 0 (0%) | 1 (1.6%) |  |
| SARS-CoV-2 variants, n(%) |  |  | 0.3 |
| alpha | 1 (2.6%) | 6 (9.7%) |  |
| delta | 7 (18%) | 6 (9.7%) |  |
| omicron | 8 (21%) | 10 (16%) |  |
| unknown | 23 (59%) | 40 (65%) |  |
| *Lab parameters* |  |  |  |
| Creatine Kinase (U/L), median IQR) | 91.0 (70.0, 111.0) | 71.0 (54.0, 92.0) | **0.009\*\*** |
| *Comorbidities* |  |  |  |
| Psychiatric Diagnosis preexisting, n(%) | 2 (5.3%) | 6 (9.7%) | 0.7 |
| Depressive diagnosis ongoing, n(%) | 9 (23%) | 10 (16%) | 0.5 |

We next compared RVA parameters across NI, recovered, and PCS patients with or without ME/CFS. AVR, CRAE, and vFID differed across groups (**Figure 4a-c**). In post-hoc analyses, vFID was significantly lower in PCS patients fulfilling ME/CFS criteria compared with recovered (p = 0.046), whereas no difference was observed relative to NI (p = 0.890). CRAE was likewise reduced in PCS patients with ME/CFS compared with recovered (p = 0.020), while the comparison with NI individuals did not reach statistical significance (p = 0.263). AVR was markedly lower in PCS patients with ME/CFS than in both NI controls (p < 0.001) and recovered individuals (p = 0.010) (**Supplementary Table 9)**.

**Figure 4.**
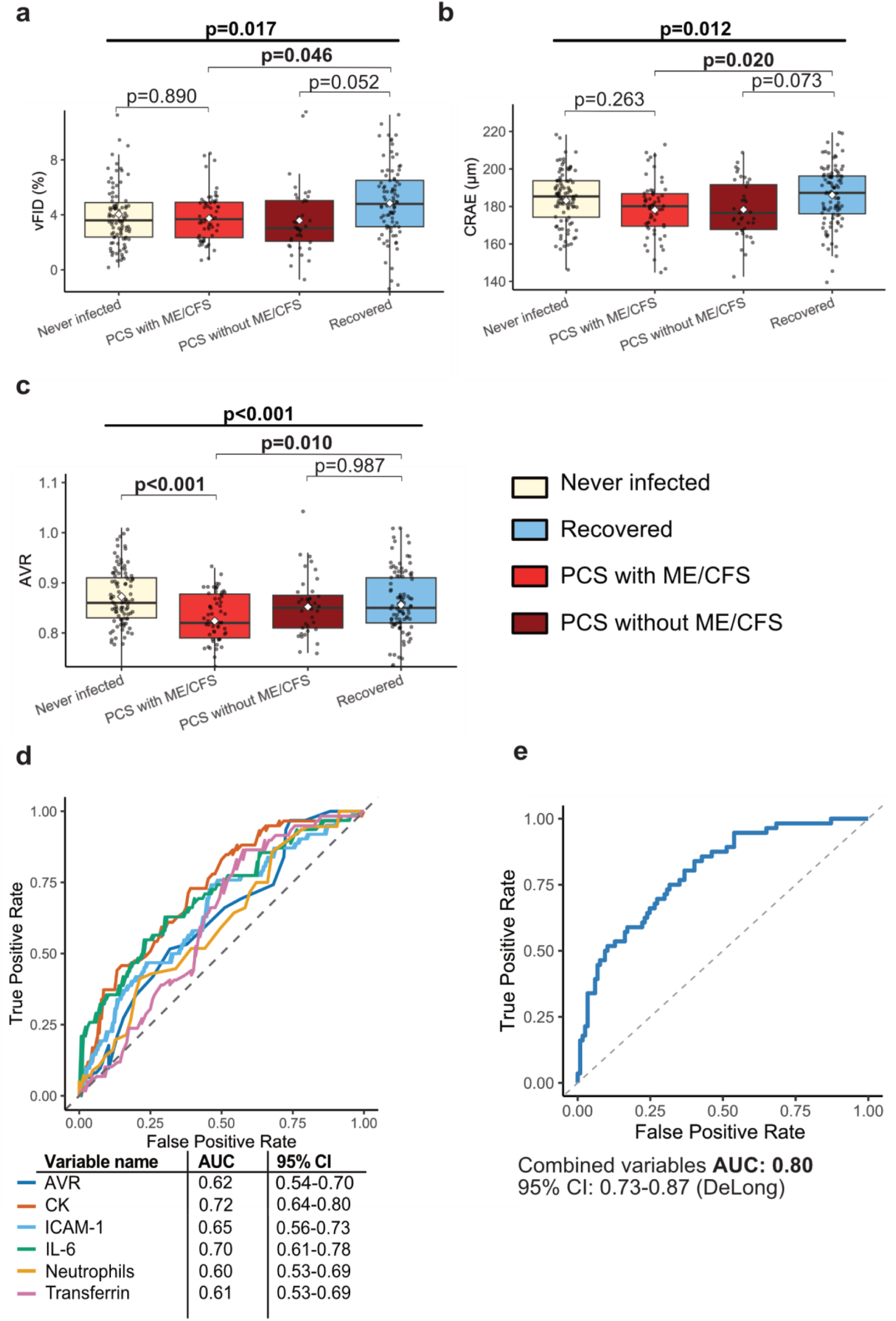
Retinal microvascular parameters and biomarker-based discrimination of ME/CFS. (a–c) Box plots of the DVA parameter vFID (a) and SVA parameters CRAE (b) and AVR (c) across four cohorts: never infected (n = 96, cornsilk), PCS with ME/CFS (n = 62, red), PCS without ME/CFS (n = 39, dark red), and recovered individuals (n = 102, light blue). Box plots indicate the median (horizontal line) and mean (black dot). Group comparisons were performed using one-way ANOVA for normally distributed variables or the Kruskal-Wallis test for non-normally distributed variables, with post-hoc testing using Tukey’s honestly significant difference (HSD) test for ANOVA models or Dunn’s test with multiplicity correction for Kruskal-Wallis models. Adjusted p-values are shown above group comparisons. Each data point represents one eye from one individual (one eye analyzed per participant). (d) Receiver operating characteristic (ROC) curves for individual biomarkers discriminating PCS patients fulfilling ME/CFS criteria from recovered individuals and PCS patients without ME/CFS. Variables include AVR (dark blue), creatine kinase (CK; red), C-reactive protein (CRP; light blue), interleukin-6 (IL-6; green), low-density lipoprotein cholesterol (LDL; orange), and transferrin (purple) (colors as indicated). (e) ROC curve showing discrimination between ME/CFS patients and the combined comparator group (recovered individuals and PCS patients without ME/CFS) using a multivariable model integrating AVR, CK, ICAM-1, IL-6, transferrin, and neutrophils. Area under the curve (AUC) with 95% confidence interval (CI) was calculated using DeLong’s method.

To determine whether an altered retinal microcirculation is specifically associated with ME/CFS, we performed multivariable regression analyses adjusting for age, sex, BMI, systolic blood pressure before RVA, and arterial hypertension. Using PCS with ME/CFS as the reference group, AVR was significantly higher in all other cohorts: NI (Std. β = 0.32, p < 0.001), PCS without ME/CFS (Std. β = 0.14, p = 0.049), and recovered individuals (Std. β = 0.17, p = 0.028). CRAE did not differ significantly after adjustment, although a trend toward wider arterioles was observed in NI individuals (Std. β = 0.14, p = 0.063) and recovered (Std. β = 0.11, p = 0.156). Recovered individuals showed significantly higher vFID compared to PCS with ME/CFS (Std. β = 0.23, p = 0.004), whereas differences between the remaining groups did not reach significance (**Supplementary Table 10**).

Finally, we performed receiver operating characteristic (ROC) analyses to identify biomarkers discriminating non-ME/CFS individuals (recovered and PCS without ME/CFS) from patients fulfilling ME/CFS criteria. Across inflammatory, iron metabolism, lipid, hematologic, and RVA parameters, 13 markers showed discriminatory performance (**Supplementary Table 11)**. From an initial panel of 13 candidate variables, six markers (CK, IL-6, ICAM-1, transferrin, AVR, neutrophils) were selected for combined ROC analysis based on individual discriminatory performance, biological plausibility, and representation of distinct pathophysiological domains (**Figure 4 d**). This integrative approach resulted in improved discrimination between ME/CFS and non-ME/CFS individuals (AUC= 0.80; 95% CI: 0.73-0.87) (**Figure 4 e**). Together, these findings indicate that PCS patients with ME/CFS exhibit the most pronounced retinal microvascular alterations, particularly reduced AVR and that these changes persist even after adjustment for relevant CV risk factors.

### Biomarkers of ED and Inflammation in Recovered, PCS and ME/CFS Patients and their Association with RVA parameters

Circulating biomarker levels were compared between recovered and PCS patients, to study potential ongoing endothelial activation and chronic inflammation. IL-6 levels were higher in PCS patients compared with recovered individuals (median 1.32 pg/mL vs. 0.92 pg/mL; p = 0.028). MCP-1 showed a non-significant trend toward higher levels in PCS (median 24.2 pg/mL vs. 18.5 pg/mL; p = 0.063). CC-Chemokine-Ligand-5 (CCL-5) did not differ between groups (p = 0.492), nor did CXCL-10 (p = 0.985) (**Figure 5 a - c, f**). Among ED markers, ICAM-1 levels were elevated in PCS compared with recovered participants (mean 3832 ± 636 pg/mL vs. 3505 ± 683 pg/mL, p <0.001). VCAM-1 was also increased in PCS (median 8073 pg/mL vs. 5899 pg/mL; p <0.001) VEGF concentrations did not differ between groups (median 6.56 pg/mL [PCS] vs. 7.68 pg/mL [recovered]; p = 0.339) (**Figure 5 d, e and g**). After adjustment for age, the associations of higher ICAM-1 (β = −358.9 pg/mL, p < 0.001) and VCAM-1 (β = −2319.6 pg/mL, p < 0.001) with PCS remained significant. In contrast, the associations with IL-6 (β = −0.24 pg/mL, p = 0.14) and MCP-1 (β = −3.52 pg/mL, p = 0.11) were attenuated (**Supplementary Table 11**). We additionally compared laboratory variables and circulating biomarkers between non-ME/CFS (recovered and PCS) with ME/CFS patients after correction for age. As shown in PCS patients, markers of ED (VCAM-1, ICAM-1) and inflammation (IL-6) were associated with ME/CFS status (**Figure 5 h**).

**Figure 5.**
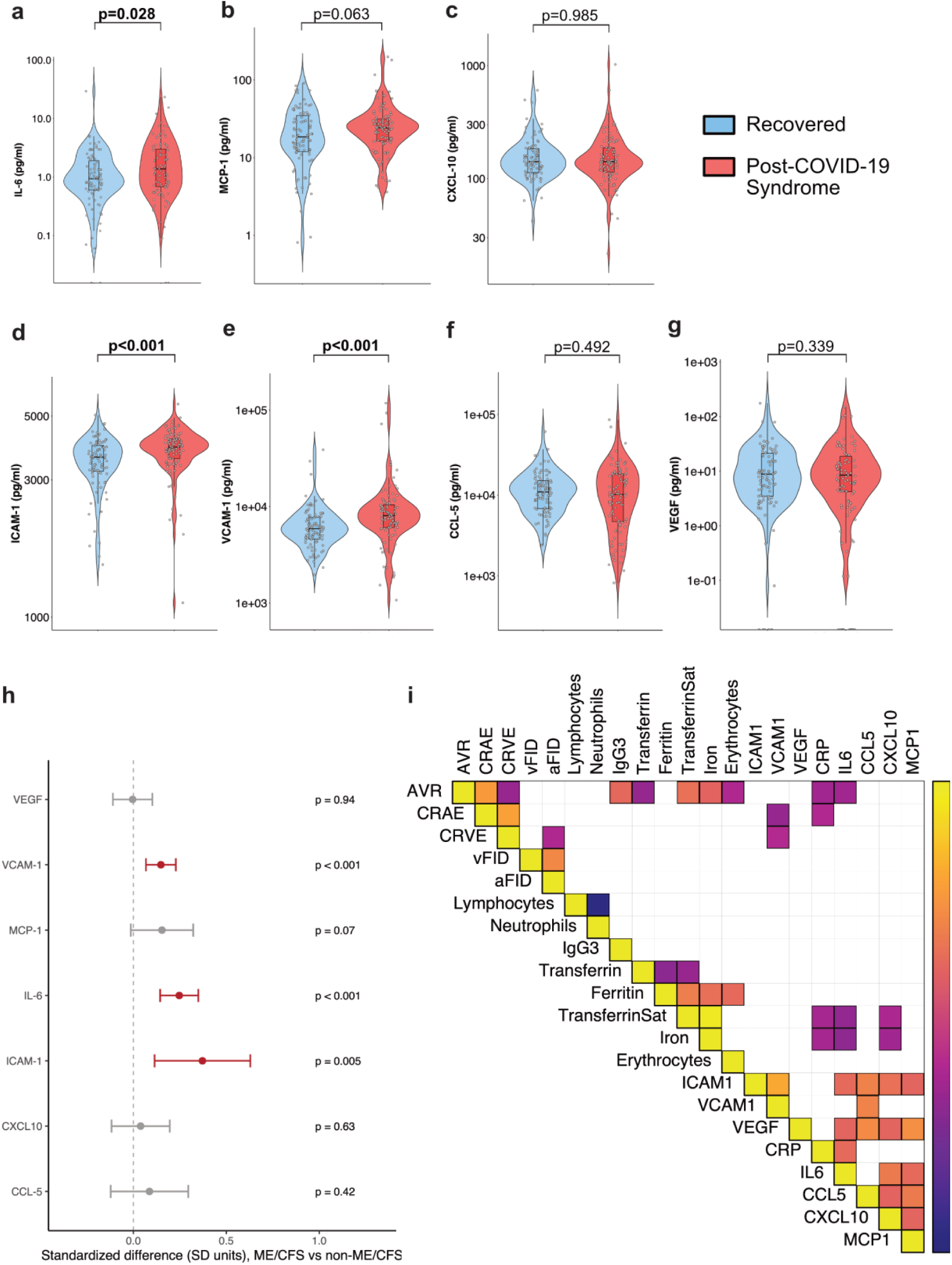
Circulating inflammatory and endothelial dysfunction markers in PCS and their associations with retinal microvascular parameters. (a–g) Violin plots illustrate circulating levels of inflammatory markers (IL-6, MCP-1, CXCL-10, CCL-5) and endothelial dysfunction markers (ICAM-1, VCAM-1, VEGF) in recovered individuals (light blue) and patients with PCS (red). Individual data points represent single participants. Box plots indicate the median (black line) and interquartile range (IQR). Group comparisons were performed using the Wilcoxon rank-sum test for non-normally distributed variables and Welch’s two-sample t-test for normally distributed variables, as appropriate. IL-6, CXCL-10, MCP-1, ICAM-1, VCAM-1, CCL-5, and VEGF were measured in n = 90 recovered individuals and n = 100 PCS patients. (h) Forest plot illustrating age-adjusted associations between ME/CFS status and seven circulating biomarkers. Effect sizes are presented as standardized differences (SD units) with 95% confidence intervals derived from regression models adjusted for age. Statistically significant associations are highlighted in red (as indicated, p<0.05). (i) Correlation heatmap showing significant Spearman correlations (p < 0.05) between selected laboratory parameters and retinal vessel analysis (RVA) parameters AVR, CRAE, CRVE, vFID, and aFID in PCS patients. Spearman’s ρ is color-coded from +1 (yellow, positive correlation) to −1 (blue, negative correlation).

We next asked whether circulating biomarkers were linked to retinal microvascular alterations within the post-infectious spectrum itself. We therefore examined the associations between circulating biomarkers and RVA parameters in all PCS patients (**Figure 5 i**). AVR showed significant inverse correlations with CRP (ρ = −0.27, p = 0.007), IL-6 (ρ = −0.27, p = 0.006), transferrin (ρ = −0.34, p <0.001), and erythrocyte count (ρ = −0.21, p = 0.037), while positive correlations were observed with IgG3 (ρ = 0.23, p = 0.020), serum iron (ρ = 0.20, p = 0.045), and transferrin saturation (ρ = 0.28, p = 0.0055). Additionally, there was a correlation for lower CRAE with higher CRP (ρ = −0.20, p = 0.045) and higher VCAM-1 (ρ = −0.33, p < 0.001), and an inverse correlation between CRVE and VCAM-1 (ρ = −0.20, p=0.047). After correction for multiple testing, the correlations between AVR and IL-6, CRP, transferrin, and transferrin saturation and the correlation between CRAE and VCAM-1 remained statistically significant (**Supplementary Figure 12**).

## Discussion

This observational case-control study is the first to perform an in-depth assessment of retinal microvascular alterations using RVA as a surrogate of persistent ED across a large, well-characterized cohort of NI individuals, recovered participants, and patients with PCS. We demonstrate that PCS patients exhibit markedly reduced venular FID, narrower retinal arterioles and a lower AVR compared with an age- and sex-matched healthy cohort. Higher symptom burden especially neurocognitive symptoms are associated with microvascular remodeling, as indicated by a lower AVR. These findings replicate and expand upon our previously published pilot study, in which comparable reductions were observed in a smaller PCS cohort compared to NI [44].

A key unresolved question was whether ED resolves during recovery or persists in association with post-viral symptomatology. Our subgroup analysis demonstrated lower vFID in PCS patients compared with recovered. These differences were independent of CV risk factors and remained evident after strict age-weighted analyses, supporting an association between persistent ED and ongoing post-viral syndromes. Reduced vFID provides functional evidence of impaired neurovascular (NV) coupling and diminished endothelial-dependent vasodilatation. Retinal FID largely depends on NO release and glia cell-regulated NV coupling [49]. In individuals recovering from an acute infection, especially in those with pre-existing CV risk or severe acute illness longitudinal studies demonstrate persistent ED, by means of impaired endothelial glycocalyx, reduced NO bioavailability and vascular function [24, 50–53]. In contrast, recovered patients without relevant comorbidities generally show only minimal or transient ED, with biomarker levels returning toward baseline over time [54–58]. Our findings suggest that patients with PCS fail to adequately restore endothelial integrity, potentially due to unresolved chronic inflammation, as reflected by persistently elevated IL-6, acute-phase proteins, and endothelial activation markers such as ICAM-1 and VCAM-1 in our cohort, which is consistent with previous reports. [7, 59, 60].

Studies in PCS report gliosis and blood-brain barrier (BBB) disruption, and sustained neuroinflammation has been linked to persistent cognitive and affective symptoms in these patients [61, 62]. Additionally, the potent vasoconstrictor endothelin-1 (ET-1), which induces neuronal apoptosis and retinal ganglion cell loss, has been shown to be elevated in PCS patients and to correlate with ongoing symptom burden. Together, these findings support NV unit dysfunction in PCS and provide a plausible explanation for the observed reduction in vFID [24, 63, 64].

Regarding SVA parameters, we observed lower AVR and CRAE in PCS compared with both recovered and NI individuals, indicating persistent microvascular remodeling (CRAE) and impaired retinal autoregulation (AVR). Retinal calibers depend on age, diabetes, BMI, and hypertension [45]. Although the association between lower AVR and PCS was attenuated after adjustment for confounders and may partly reflect the higher CV risk profile of PCS patients compared with recovered, the parallel associations of both AVR and arteriolar FID with clinical severity suggest that retinal ED is not solely attributable to baseline risk factors but is also linked to symptom burden.

Individuals with predominantly ongoing neurocognitive symptoms showed a lower AVR than recovered individuals or PCS patients without these symptoms. Given the established associations of lower AVR and arteriolar narrowing with cerebral small vessel disease, neuroinflammation, and cognitive impairment, reduced AVR and CRAE observed in PCS provide evidence of persistent microvascular injury affecting neurovascular integrity and offer a plausible mechanistic link to ongoing neurocognitive symptoms [65–68]. Altered SVA parameters in general have been consistently linked to CV risk in landmark studies. Independently, epidemiological data demonstrate increased long-term CV risk after acute COVID-19 infection [21, 22, 31, 32]. In this context, our observation of narrow retinal arterioles and lower AVR in PCS indicates sustained microvascular remodeling beyond the acute phase of infection. Given the median disease duration of 13 months in our cohort, the persistence of these microvascular alterations suggests that RVA may provide a non-invasive window into sustained microvascular injury and potentially increased CV risk in patients with PCS.

Early recognition of the clinical and biological overlap between ME/CFS and PCS has shifted research toward a unified post-infectious disease framework [7, 15]. Accordingly, 61% of PCS patients in our cohort met ME/CFS criteria and showed markedly greater symptom burden and functional impairment.

When examining all previously infected individuals, we observed a continuous gradient of retinal ED, with progressively lower vFID, CRAE, and AVR values across increasing clinical severity. PCS patients meeting ME/CFS criteria showed the most pronounced impairments, suggesting that retinal ED reflects the severity of the post-infectious phenotype.

After adjustment for age, IL-6, ICAM-1, and VCAM-1 remained elevated in patients with ME/CFS. Notably, a similar biomarker signature was present in PCS more broadly, suggesting a shared inflammatory-endothelial activation axis across the post-infectious spectrum, with ME/CFS representing a clinically more severe or biologically enriched phenotype [19, 63, 69].

AVR alone provided modest discrimination between ME/CFS and non-ME/CFS patients, however its combination with routine laboratory parameters substantially improved accuracy reaching an AUC of 0.80 comparable to a previously published diagnostic cell-based blood test for ME/CFS [70, 71]. These findings suggest that monitoring endothelial health via RVA may serve as a valuable adjunct for biological stratification in patients with ongoing post-viral symptoms in general.

Lower AVR was correlated with higher IL-6 and CRP, linking retinal microvascular remodeling to ongoing systemic inflammation. Both IL-6 and CRP have been implicated in persistent neurocognitive symptoms in PCS, and elevated IL-6 during acute infection predicts worse long-term cognitive performance [72–75]. IL-6-mediated neuroinflammation, including BBB disruption and impaired NV coupling, provides a plausible mechanism linking inflammation to reduced AVR [76]. Narrower retinal arterioles were additionally associated with higher VCAM-1 levels, further supporting an inflammation-driven ED phenotype.

Beyond inflammatory signaling, IL-6 is a central mediator of hepcidin induction and inflammation-driven alterations in iron homeostasis [77, 78]. Consistently, IL-6 was negatively correlated with iron-related parameters in our cohort, and AVR was associated with transferrin and transferrin saturation. These findings suggest that retinal microvascular alterations in PCS co-occur with an inflammatory–iron axis rather than isolated iron deficiency. Persistent iron homeostasis dysregulation and inflammatory stress erythropoiesis have been reported following COVID-19 and linked to long-term PCS outcomes [14, 77]. Finally, lower IgG3 levels, previously associated with an increased risk of PCS, were confirmed in our cohort and correlated inversely with AVR, suggesting that immune dysregulation may further contribute to microvascular alterations in PCS [79].

### Limitations

Several limitations warrant consideration. First, this is a cross-sectional, single-center cohort study; therefore, the observed associations, while biologically plausible, should not be interpreted as causal. The present findings are exploratory and hypothesis-generating and require validation in larger, independent cohorts. Second, despite adjustment for potential confounders, the recovered cohort was significantly younger than the PCS group, which may have contributed to differences in retinal microvascular parameters. However, altered RVA measures were also observed when PCS patients were compared with the NI cohort, which was older and had a higher burden of comorbidities, supporting the robustness of the findings. Finally, the cross-sectional design precludes conclusions regarding whether PCS patients with more severe symptoms and adverse retinal microvascular phenotypes are at increased CV risk. Prospective longitudinal studies integrating RVA with adjudicated CV outcomes are required to determine whether retinal microvascular measures can identify a high-risk PCS subpopulation that may benefit from intensified CV monitoring and preventive strategies.

## Conclusion

This study demonstrates that patients with PCS exhibit persistent functional and structural alterations of the retinal microvasculature, indicating ongoing endothelial and neurovascular dysfunction. Retinal ED is most pronounced in patients with higher symptom burden and in those fulfilling ME/CFS criteria, and is accompanied by sustained inflammatory and endothelial activation, as reflected by elevated IL-6, ICAM-1, and VCAM-1. The observed associations between retinal microvascular parameters, systemic inflammation, and iron homeostasis suggest that microvascular remodeling in PCS reflects a broader, inflammation-driven vascular phenotype rather than isolated CV risk. Together, these findings support the utility of RVA as a non-invasive window into post-infectious ED and highlight its potential role in biological stratification and risk assessment in patients with persistent symptoms. Prospective longitudinal studies are warranted to determine the prognostic value of retinal microvascular measures and their relevance for long-term CV outcomes in this population.

## Data Availability

The datasets used during the current study are available from the corresponding author on reasonable request.

## Abbreviations

aFID: Arteriolar flicker-induced dilation
ANOVA: Analysis of variance
ATO: Average treatment effect in the overlap population
AVR: Arteriolar-to-venular ratio
BBB: Blood–brain barrier
BMI: Body mass index
CCL-5: CC-Chemokine-Ligand-5
CK: Creatine kinase
CRAE: Central retinal artery equivalent
CRP: C-reactive protein
CRVE: Central retinal vein equivalent
CV: Cardiovascular
C19-YRS: COVID-19 Yorkshire Rehabilitation Scale
CXCL10 (IP-10): C-X-C motif chemokine 10 (Interferon gamma-induced protein-10)
DVA: Dynamic retinal vessel analysis
ED: Endothelial dysfunction
EQ-5D-5L: EuroQol five-dimension five-level questionnaire
FID: Flicker-induced dilation
FMD: Flow-mediated dilation
FSS: Fatigue Severity Scale
GAD-7: Generalized Anxiety Disorder-7
HC: Healthy controls
HIF: Hypoxia-inducible factor
ICAM-1: Intercellular adhesion molecule-1
ICTRP: International Clinical Trials Registry Platform
IgG: Immunoglobulin G
IL-6: Interleukin-6
ME/CFS: Myalgic encephalomyelitis/chronic fatigue syndrome
MCP-1: Monocyte chemoattractant protein-1
NI: Never infected
NO: Nitric oxide
NV: Neurovascular
OCT-A: Optical coherence tomography angiography
PCS: Post-COVID-19 syndrome
PEM: Post-exertional malaise
PHQ-9: Patient Health Questionnaire-9
POH: Post-occlusive reactive hyperemia
PROMs: Patient-reported outcome measures
PWV: Pulse-wave velocity
ROC: Receiver operating characteristic
RVA: Retinal vessel analysis
SD: Standard deviation
SVA: Static retinal vessel analysis
VCAM-1: Vascular cell adhesion molecule-1
VEGF: Vascular endothelial growth factor
vFID: Venular flicker-induced dilation
WHO: World Health Organization

## Acknowledgements

We want to thank all patients and individuals willing to take part in this study. Additionally, we want to thank the doctoral students, mainly A. Woehnle, V.Kessler and J. Negele involved.

## Contribution

CS, the principal investigator, and TW conceived the study and led the project. MW contributed to data interpretation and analysis, and drafting of the manuscript. ML, IL, and AR served as methodologists for the basic science components and bio sample processing and analyses. RG contributed to the development of the study and the manuscript. BH assisted with figure creation and statistical and data analyses. HH, LS, provided data on the healthy cohort and expertise in RVA. FCO, and LGS provided significant expertise in RVA. All authors contributed to the manuscript’s development and ensured the accuracy of all parts of the work. All authors read and approved the final version.

## Corresponding author

Timon Wallraven is the corresponding author.

## Ethics declaration

### Ethics approval and consent to participate

The study protocol was approved by the Ethics Committee of the Technical University of Munich, School of Medicine, Klinikum rechts der Isar (Approval number: 2022-317-S-SR) and the Ethics Committee of North-western and Central Switzerland (EKNZ 2017-01451). This study is registered at ClinicalTrials.gov (NCT05635552).

### Consent for publication

Not applicable

## References

1. Lopez-Leon, S., et al., More Than 50 Long-Term Effects of COVID-19: A Systematic Review and Meta-Analysis. Res Sq, 2021.

2. Lai, C.C., et al., Long COVID: An inevitable sequela of SARS-CoV-2 infection. J Microbiol Immunol Infect, 2023. 56(1): p. 1–9.

3. Fernandez-de-Las-Peñas, C., et al., Persistence of post-COVID symptoms in the general population two years after SARS-CoV-2 infection: A systematic review and meta-analysis. J Infect, 2024. 88(2): p. 77–88.

4. Scott, A., et al., Substantial health and economic burden of COVID-19 during the year after acute illness among US adults not at high risk of severe COVID-19. BMC Medicine, 2024. 22(1): p. 47.

5. Soriano, J.B., et al., A clinical case definition of post-COVID-19 condition by a Delphi consensus. Lancet Infect Dis, 2022. 22(4): p. e102–e107.

6. Greenhalgh, T., et al., Long COVID: a clinical update. Lancet, 2024. 404(10453): p. 707–724.

7. Kedor, C., et al., A prospective observational study of post-COVID-19 chronic fatigue syndrome following the first pandemic wave in Germany and biomarkers associated with symptom severity. Nature Communications, 2022. 13(1): p. 5104.

8. Vernon, S.D., et al., Incidence and Prevalence of Post-COVID-19 Myalgic Encephalomyelitis: A Report from the Observational RECOVER-Adult Study. J Gen Intern Med, 2025. 40(5): p. 1085–1094.

9. Østergaard, L., SARS CoV-2 related microvascular damage and symptoms during and after COVID-19: Consequences of capillary transit-time changes, tissue hypoxia and inflammation. Physiol Rep, 2021. 9(3): p. e14726.

10. Owens, C.D., et al., COVID-19 Exacerbates Neurovascular Uncoupling and Contributes to Endothelial Dysfunction in Patients with Mild Cognitive Impairment. Biomolecules, 2024. 14(12).

11. Xu, S.W., I. Ilyas, and J.P. Weng, Endothelial dysfunction in COVID-19: an overview of evidence, biomarkers, mechanisms and potential therapies. Acta Pharmacol Sin, 2023. 44(4): p. 695–709.

12. Wu, X., et al., Damage to endothelial barriers and its contribution to long COVID. Angiogenesis, 2024. 27(1): p. 5–22.

13. Bonaventura, A., et al., Endothelial dysfunction and immunothrombosis as key pathogenic mechanisms in COVID-19. Nature Reviews Immunology, 2021. 21(5): p. 319–329.

14. Gupta, G., et al., Mechanistic Insights Into Long Covid: Viral Persistence, Immune Dysregulation, and Multi-Organ Dysfunction. Compr Physiol, 2025. 15(3): p. e70019.

15. Choutka, J., et al., Unexplained post-acute infection syndromes. Nature Medicine, 2022. 28(5): p. 911–923.

16. McLaughlin, M., et al., People with Long COVID and Myalgic Encephalomyelitis/Chronic Fatigue Syndrome Exhibit Similarly Impaired Vascular Function. Am J Med, 2025. 138(3): p. 560–566.

17. Fan, B.E., et al., Hypercoagulability, endotheliopathy, and inflammation approximating 1 year after recovery: Assessing the long-term outcomes in COVID-19 patients. Am J Hematol, 2022. 97(7): p. 915–923.

18. Chioh, F.W., et al., Convalescent COVID-19 patients are susceptible to endothelial dysfunction due to persistent immune activation. Elife, 2021. 10.

19. Maksoud, R., et al., Biomarkers for myalgic encephalomyelitis/chronic fatigue syndrome (ME/CFS): a systematic review. BMC Med, 2023. 21(1): p. 189.

20. Domingo, J.C., et al., Association of circulating biomarkers with illness severity measures differentiates myalgic encephalomyelitis/chronic fatigue syndrome and post-COVID-19 condition: a prospective pilot cohort study. J Transl Med, 2024. 22(1): p. 343.

21. Bruno, R.M., et al., Accelerated vascular ageing after COVID-19 infection: the CARTESIAN study. European Heart Journal, 2025.

22. Battistoni, A., et al., Persistent increase of cardiovascular and cerebrovascular events in COVID-19 patients: a 3-year population-based analysis. Cardiovasc Res, 2024. 120(6): p. 623–629.

23. Xie, Y., et al., Long-term cardiovascular outcomes of COVID-19. Nature Medicine, 2022. 28(3): p. 583–590.

24. Haffke, M., et al., Endothelial dysfunction and altered endothelial biomarkers in patients with post-COVID-19 syndrome and chronic fatigue syndrome (ME/CFS). Journal of Translational Medicine, 2022. 20(1): p. 138.

25. Ståhlberg, M., et al., Post-Acute COVID-19 Syndrome: Prevalence of Peripheral Microvascular Endothelial Dysfunction and Associations With NT-ProBNP Dynamics. Am J Med, 2025. 138(6): p. 1019–1028.

26. Mavraganis, G., et al., Clinical implications of vascular dysfunction in acute and convalescent COVID-19: A systematic review. Eur J Clin Invest, 2022. 52(11): p. e13859.

27. Hosp, J.A., et al., Cerebral microstructural alterations in Post-COVID-condition are related to cognitive impairment, olfactory dysfunction and fatigue. Nat Commun, 2024. 15(1): p. 4256.

28. Shah, W., et al., Exploring Endothelial Cell Dysfunction’s Impact on the Brain-Retina Microenvironment Connection: Molecular Mechanisms and Implications. Mol Neurobiol, 2025. 62(6): p. 7484–7505.

29. Hanssen, H., L. Streese, and W. Vilser, Retinal vessel diameters and function in cardiovascular risk and disease. Progress in Retinal and Eye Research, 2022. 91: p. 101095.

30. Gunthner, R., et al., Endothelial dysfunction in retinal vessels of hemodialysis patients compared to healthy controls. Sci Rep, 2024. 14(1): p. 13948.

31. Seidelmann, S.B., et al., Retinal Vessel Calibers in Predicting Long-Term Cardiovascular Outcomes: The Atherosclerosis Risk in Communities Study. Circulation, 2016. 134(18): p. 1328–1338.

32. Yatsuya, H., et al., Retinal microvascular abnormalities and risk of lacunar stroke: Atherosclerosis Risk in Communities Study. Stroke, 2010. 41(7): p. 1349–55.

33. Takayanagi, Y., et al., Association between Systemic Antioxidant Capacity and Retinal Vessel Diameters in Patients with Primary-Open Angle Glaucoma. Life (Basel), 2020. 10(12).

34. Günthner, R., et al., Mortality prediction of retinal vessel diameters and function in a long-term follow-up of haemodialysis patients. Cardiovascular Research, 2022. 118(16): p. 3239–3249.

35. van Dinther, M., et al., Retinal microvascular function is associated with the cerebral microcirculation as determined by intravoxel incoherent motion MRI. J Neurol Sci, 2022. 440: p. 120359.

36. Kuchler, T., et al., All eyes on PCS: analysis of the retinal microvasculature in patients with post-COVID syndrome-study protocol of a 1 year prospective case-control study. Eur Arch Psychiatry Clin Neurosci, 2023.

37. Bahmer, T., et al., Severity, predictors and clinical correlates of Post-COVID syndrome (PCS) in Germany: A prospective, multi-centre, population-based cohort study. EClinicalMedicine, 2022. 51: p. 101549.

38. O’Connor, R.J., et al., The COVID-19 Yorkshire Rehabilitation Scale (C19-YRS): Application and psychometric analysis in a post-COVID-19 syndrome cohort. J Med Virol, 2022. 94(3): p. 1027–1034.

39. Krupp, L.B., et al., Fatigue in multiple sclerosis. Arch Neurol, 1988. 45(4): p. 435–7.

40. Costantini, L., et al., Screening for depression in primary care with Patient Health Questionnaire-9 (PHQ-9): A systematic review. J Affect Disord, 2021. 279: p. 473–483.

41. Spitzer, R.L., et al., A brief measure for assessing generalized anxiety disorder: the GAD-7. Arch Intern Med, 2006. 166(10): p. 1092–7.

42. Herdman, M., et al., Development and preliminary testing of the new five-level version of EQ-5D (EQ-5D-5L). Qual Life Res, 2011. 20(10): p. 1727–36.

43. Encephalomyelitis/Chronic, C.o.t.D.C.f.M., et al., Beyond Myalgic Encephalomyelitis/Chronic Fatigue Syndrome: Redefining an Illness. 2015.

44. Kuchler, T., et al., Correction: Persistent endothelial dysfunction in post-COVID-19 syndrome and its associations with symptom severity and chronic inflammation. Angiogenesis, 2023. 26(4): p. 565.

45. Nagel, E., W. Vilser, and I. Lanzl, Age, blood pressure, and vessel diameter as factors influencing the arterial retinal flicker response. Invest Ophthalmol Vis Sci, 2004. 45(5): p. 1486–92.

46. Kotliar, K.E., et al., Dynamic retinal vessel response to flicker in obesity: A methodological approach. Microvasc Res, 2011. 81(1): p. 123–8.

47. Ponto, K.A., et al., Retinal vessel metrics: normative data and their use in systemic hypertension: results from the Gutenberg Health Study. J Hypertens, 2017. 35(8): p. 1635–1645.

48. Yuen, V.L., et al., Effects of firsthand tobacco smoking on retinal vessel caliber: a systematic review and meta-analysis. Graefes Arch Clin Exp Ophthalmol, 2024. 262(5): p. 1397–1407.

49. Hanssen, H., L. Streese, and W. Vilser, Retinal vessel diameters and function in cardiovascular risk and disease. Prog Retin Eye Res, 2022. 91: p. 101095.

50. Charfeddine, S., et al., *c*. Front Cardiovasc Med, 2021. 8: p. 745758.

51. Lambadiari, V., et al., Association of COVID-19 with impaired endothelial glycocalyx, vascular function and myocardial deformation 4 months after infection. Eur J Heart Fail, 2021. 23(11): p. 1916–1926.

52. Dominic, P., et al., Decreased availability of nitric oxide and hydrogen sulfide is a hallmark of COVID-19. Redox Biol, 2021. 43: p. 101982.

53. Lip, S., et al., Long-term effects of SARS-CoV-2 infection on blood vessels and blood pressure - LOCHINVAR. J Hypertens, 2025. 43(6): p. 1057–1065.

54. Kozłowski, P., et al., Mild-to-Moderate COVID-19 Convalescents May Present Pro-Longed Endothelium Injury. J Clin Med, 2022. 11(21).

55. Oikonomou, E., et al., Endothelial dysfunction in acute and long standing COVID-19: A prospective cohort study. Vascul Pharmacol, 2022. 144: p. 106975.

56. Ambrosino, P., et al., Clinical assessment of endothelial function in convalescent COVID-19 patients: a meta-analysis with meta-regressions. Annals of Medicine, 2022. 54(1): p. 3233–3248.

57. Zanoli, L., et al., Vascular Dysfunction of COVID-19 Is Partially Reverted in the Long-Term. Circulation Research, 2022. 130(9): p. 1276–1285.

58. Zhang, C., et al., New findings on retinal microvascular changes in patients with primary COVID-19 infection: a longitudinal study. Front Immunol, 2024. 15: p. 1404785.

59. Greene, C., et al., Author Correction: Blood-brain barrier disruption and sustained systemic inflammation in individuals with long COVID-associated cognitive impairment. Nat Neurosci, 2024. 27(5): p. 1019.

60. Kedor, C., et al., A prospective observational study of post-COVID-19 chronic fatigue syndrome following the first pandemic wave in Germany and biomarkers associated with symptom severity. Nature Communications, 2022. 13(1): p. 5104.

61. Braga, J., et al., Neuroinflammation After COVID-19 With Persistent Depressive and Cognitive Symptoms. JAMA Psychiatry, 2023. 80(8): p. 787–795.

62. Greene, C., et al., Blood-brain barrier disruption and sustained systemic inflammation in individuals with long COVID-associated cognitive impairment. Nat Neurosci, 2024. 27(3): p. 421–432.

63. Jacobs, L.M.C., et al., Biomarkers of sustained systemic inflammation and microvascular dysfunction associated with post-COVID-19 condition symptoms at 24 months after SARS-CoV-2-infection. Front Immunol, 2023. 14: p. 1182182.

64. Prasanna, G., R. Krishnamoorthy, and T. Yorio, Endothelin, astrocytes and glaucoma. Exp Eye Res, 2011. 93(2): p. 170–7.

65. de Jong, F.J., et al., Retinal vascular caliber and risk of dementia: the Rotterdam study. Neurology, 2011. 76(9): p. 816–21.

66. Arnould, L., et al., Retinal Microvascular Correlates of Cerebral Small Vessel Disease in Older Age. Neurology, 2025. 105(9): p. e214251.

67. Jiang, K., et al., Midlife Retinal Microvascular Signs and Late-Life Neuroimaging Features of Cerebral Small Vessel Disease in the ARIC Study. Neurology, 2025. 105(4): p. e213919.

68. Luyten, L.J., et al., Association of Retinal Microvascular Characteristics With Short-term Memory Performance in Children Aged 4 to 5 Years. JAMA Netw Open, 2020. 3(7): p. e2011537.

69. Russell, L., et al., Illness progression in chronic fatigue syndrome: a shifting immune baseline. BMC Immunol, 2016. 17: p. 3.

70. Missailidis, D., et al., Cell-Based Blood Biomarkers for Myalgic Encephalomyelitis/Chronic Fatigue Syndrome. Int J Mol Sci, 2020. 21(3).

71. Hunter, E., et al., Development and validation of blood-based diagnostic biomarkers for Myalgic Encephalomyelitis/Chronic Fatigue Syndrome (ME/CFS) using EpiSwitch(®) 3-dimensional genomic regulatory immuno-genetic profiling. J Transl Med, 2025. 23(1): p. 1048.

72. Ferrando, S.J., et al., Associations of elevated pro-inflammatory cytokines Interleukin-6, C-reactive protein and tumor necrosis factor alpha with neuropsychiatric symptoms of post-acute sequelae of COVID-19 (PASC). J Psychiatr Res, 2025. 190: p. 128–136.

73. Nuber-Champier, A., et al., Systemic cytokines related to memory function 6-9 months and 12-15 months after SARS-CoV-2 infection. Sci Rep, 2024. 14(1): p. 22660.

74. Tilikete, C., et al., Exploring the landscape of symptom-specific inflammatory cytokines in post-COVID syndrome patients. BMC Infect Dis, 2024. 24(1): p. 1337.

75. Ogando, N.S., et al., Immunometabolism perturbations in post-COVID-19 condition: interleukin-6 and monoamine oxidase interactions drive neuropsychiatric syndromes. Brain, Behavior, and Immunity, 2025. 129: p. 690–708.

76. García-Juárez, M. and A. Camacho-Morales, Defining the Role of Anti- and Pro-inflammatory Outcomes of Interleukin-6 in Mental Health. Neuroscience, 2022. 492: p. 32–46.

77. Hanson, A.L., et al., Iron dysregulation and inflammatory stress erythropoiesis associates with long-term outcome of COVID-19. Nature Immunology, 2024. 25(3): p. 471–482.

78. Lefebvre, T., et al., Evaluation of iron metabolism in hospitalized COVID-19 patients. Clin Chim Acta, 2023. 548: p. 117509.

79. Cervia, C., et al., Immunoglobulin signature predicts risk of post-acute COVID-19 syndrome. Nat Commun, 2022. 13(1): p. 446.

